# Homicide in Pregnant and Postpartum versus Nonpregnant and Nonpostpartum Populations: Re-estimation of a Rate Ratio using a Person-time Framework

**DOI:** 10.64898/2026.01.25.26344756

**Authors:** Claire R. McNellan, Neal Marquez, Monica Alexander

**Affiliations:** Department of Health Systems, Management & Policy, Colorado School of Public Health, University of Colorado Anschutz Medical Campus, Aurora, Colorado; Population Research Center, Portland State University, Portland, Oregon; Colorado State Demography Office, Colorado Department of Local Affairs, Denver, Colorado; Department of Statistical Sciences, University of Toronto, Toronto, Ontario; Department of Sociology, University of Toronto, Toronto, Ontario

## Abstract

We aim to re-estimate the national homicide rate ratio between nonpregnant/nonpostpartum and pregnant/postpartum women accounting for person-time exposure, which prior studies overlooked. Using a theoretical framework for descriptive epidemiology, we complete a retrospective analysis to estimate the pregnancy-associated homicide rate and re-estimate the national homicide rate ratio between pregnant/postpartum and nonpregnant/nonpostpartum populations in 2020. We use National Vital Statistics System death, fetal death, birth, and Census Bureau data to identify the population at risk. We compare mortality rates and 95% confidence intervals overall and stratified by race, ethnicity, and age. Among the 9,905,908 pregnancies contributing person-time, there were 185 homicides. The relative homicide risk was 35% higher among nonpregnant/nonpostpartum compared to pregnant/postpartum populations. Pregnancy was only associated with elevated risk among ages 10-19 (homicide rate ratio 3.82; 95% CI 2.39-5.77). Homicide rate ratios between nonpregnant/nonpostpartum and pregnant/postpartum women calculated accounting for exposure time and pregnancy transitions contradict previous estimates. Accurate assessment of mortality rates is essential to develop strategies protective against maternal mortality.

## Introduction

The risk of mortality for pregnant and postpartum women in the United States is growing and heightened compared to other Organization for Economic Co-Operation and Development (OECD) countries.^1–4^ The majority of research on this topic relies on a *maternal mortality ratio*– the number of maternal deaths per 100,000 live births, where maternal deaths include those strictly due to causes related to or aggravated by pregnancy or childbirth that occur while pregnant or within 42 days of the termination of pregnancy. This research may underestimate the risks associated with pregnancy by omitting sizable non-obstetric causes, such as accidents resulting in placental abruption, suicide of women experiencing postpartum depression, or intimate partner violence^.5^ The *pregnancy-associated mortality ratio*, on the other hand, is inclusive of deaths that occur during pregnancy or up to one year after the termination of pregnancy, including deaths due to social, environmental, and non-biological factors.^6^ This indicator is of critical public health importance because it expands our understanding of the risks women face during pregnancy and postpartum periods. Such studies can aid in the development of strategies for mitigating a broader array of urgent health and safety concerns during and immediately after pregnancy such as homicide, suicide, and accidents.

The United States recently advanced public health surveillance of maternal health and safety by adding a pregnancy checkbox to the 2003 revision of the U.S. Standard Certificate of Death–a revision adopted by vital registration systems nationwide in 2018.^7^ The pregnancy checkbox can be used to identify whether a given decedent was pregnant at the time of death, pregnant within 42 days of their death, or pregnant within 1 year of their death.^6^ Estimates made using the pregnancy checkbox data reveal that the United States not only exhibits higher maternal mortality ratios compared to other OECD countries, but also higher pregnancy-associated mortality ratios for subsets of causes such as homicide^.8,9^

The availability of pregnancy information on death certificates allows for the mortality risk among pregnant/postpartum women to be compared to the mortality risk of nonpregnant/nonpostpartum women. For example, Wallace et al., 2021 estimated the first national rate ratio of homicide between pregnant/postpartum and nonpregnant/nonpostpartum populations and found that pregnancy and the postpartum period are times of elevated homicide risk.^9^ To answer such questions using vital statistics records, however, it is necessary to identify an appropriate population at risk.

Previous studies calculated a pregnancy-associated mortality ratio using the standard denominator for maternal mortality ratio: live births^.9–13^ To calculate a comparable statistic for the nonpregnant/nonpostpartum population, a denominator of the total female population minus live births was used. While using live births is appropriate for studies that focus solely on pregnant or postpartum populations, studies which evaluate the rate ratio of mortality between pregnant/postpartum and nonpregnant/nonpostpartum populations necessitate a comparable denominator that is appropriate for both populations. We argue that a pregnancy-associated mortality ratio, used in previous studies, to compare mortality between pregnant/postpartum and nonpregnant/nonpostpartum populations is inappropriate as it does not accurately represent time and the transition of women between pregnant/postpartum and nonpregnant/nonpostpartum periods. Instead, the calculation of a mortality rate is necessary because it captures the time at risk in either the pregnant/postpartum or nonpregnant/nonpostpartum periods.

In this paper, we draw upon a theoretical framework for descriptive epidemiology to outline a person-time methodology for more accurate comparison between nonpregnant/nonpostpartum and pregnant/postpartum women with respect to their homicide rates^.14^ This framework posits that the quantification of some feature of the health of a population must capture the target population anchored in time.^14^ While previous literature estimating pregnancy-associated mortality used live births as a denominator,^9–13^ this denominator fails to capture the exposure time associated with pregnancy-associated mortality. Our study uses a person-time methodology to complete a retrospective re-estimation of the national homicide rate ratio between pregnant/postpartum and nonpregnant/nonpostpartum populations in 2020. We focus solely on homicides in 2020 to mirror a previous study of pregnancy-associated mortality risk leveraging our newly presented methodology.

## Methods

Data for this retrospective analysis of 2020 mortality came from four sources. First, the total study population, women aged 10-44 in 2020 in the United States, was taken from the United States Census Bureau vintage 2024 population estimates. The 2020 mid-year population was multiplied by 12 to convert the study population to a person-month metric. The incorporation of time in our novel approach enables us to better compare mortality rates between the pregnant/postpartum and the nonpregnant/nonpostpartum population. In this way, our methodology allows for the calculation of a pregnancy-associated homicide rate rather than a pregnancy homicide ratio. The population was further divided into age groups (i.e., 10-19, 20-24, 25-29, 30-34, and 35-44), as well as by race and ethnicity (i.e., non-Hispanic (NH) White, NH Black, and Hispanic), aligning with the stratification used in previous studies.^15^

Information on deaths came from the restricted-use National Vital Statistics System (NVSS) mortality micro-data from the National Center for Health Statistics. The NVSS mortality data contain all deaths that occurred in the United States and are associated with a death certificate. The analysis was restricted to homicide deaths occurring among females aged 10-44 during the year 2020, to mirror previous studies.^15^ Homicide deaths were identified as deaths that were coded with an International Classification of Diseases, 10th Revision (ICD-10) code of X85-Y09.^16^ We identified pregnant and postpartum individuals (i.e., pregnancy-associated status) using the pregnancy checkbox data. Nonpregnant, nonpostpartum individuals were those records on which the pregnancy checkbox reflected anything other than pregnant, pregnant within 42 days, or pregnant within 1 year.

While the study population was identified using Census Bureau data, pregnancy-associated status could not be obtained from this source. To quantify the population-time at risk (i.e., identify the number of pregnant individuals and the period in which individuals were pregnant or postpartum), we used a combination of four data sources: NVSS birth, fetal death, and death records, as well as Census Bureau Vintage 2024 population data.

First, NVSS birth data were used to identify a subset of the population who experienced pregnancy-associated status during 2020. Similar to NVSS death micro-data, NVSS birth micro-data includes all births occurring in the United States. All birth records report the year and month of birth, gestation time, occurrence of multiple gestation pregnancies (e.g., twins, triplets), and maternal demographics. For each record, we calculated the gestation time in months and derived the number of pregnancy months that occurred in 2020. For example, if a birth occurred in February of 2021 with a gestation time of 10 months, 1 month of gestation time was attributed to January of 2021, and thus excluded from our analysis, while 9 months of gestation occurred in 2020 and were therefore included in our analysis. In addition, each birth was associated with 12 postpartum months beginning with the month of birth, and all postpartum months occurring in 2020 were included in our analysis. For example, if a birth occurred in July 2019, 6 months of postpartum pregnancy-associated time would be included in our analysis. No pregnancy could be associated with more than 12 months of pregnancy-associated person time for 2020. A visual example of the relationship between births, pregnancy, and postpartum periods is shown in Figure 1.

**Figure 1.**
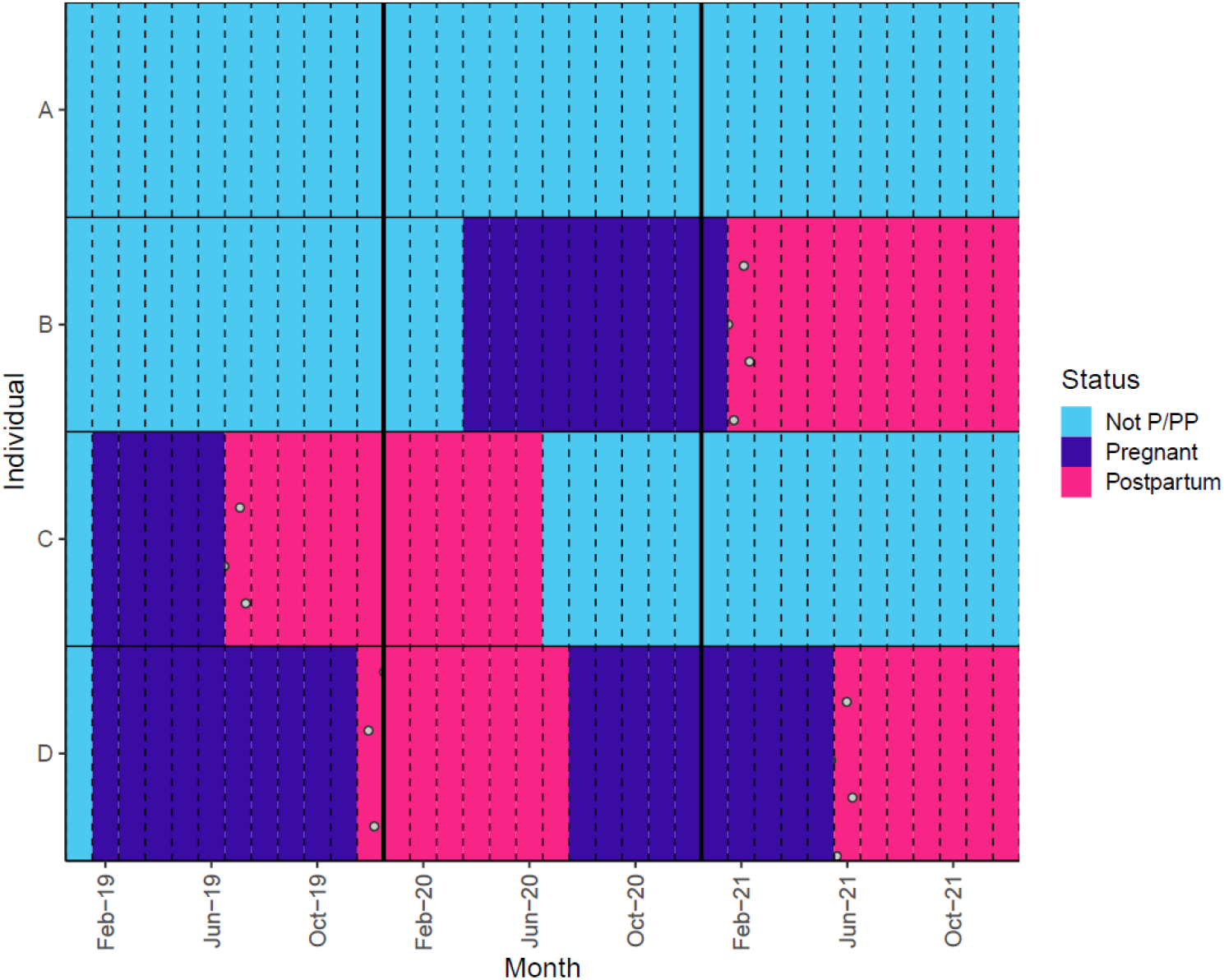
Visual representation of births and relationship to pregnancy and postpartum exposure time for 4 individuals, shown across 36 months of observation spanning the years 2019-2021. Note: Each cell represents a person-month where a mutually exclusive status of nonpregnant/nonpostpartum (Not P/PP), pregnant, or postpartum is assigned. Transitions from pregnant to postpartum represent a birth (filled with a dotted pattern). Individual A was never pregnant during the years 2019 through 2021 and thus no exposure time was recorded. Individual B had a birth in February of 2021 with 10 months of gestation time and thus 9 months of exposure time were recorded. Individual C had a birth in July of 2019 with some postpartum exposure time in 2020. Individual D had births in December of 2019 and June of 2021, with both postpartum and pregnant months contributing exposure time in 2020.

To avoid double-counting pregnancy time in multiple gestation births, a subset of records was removed from our analysis. For any birth records indicated to be part of a multiple gestation birth, records were matched by month and gestation time and deduplicated such that only one record from the multiple gestation birth contributed to pregnancy-associated time in our analysis. In other words, if a birth record with a twin was observed, another birth record with a twin with matching birth month and gestation time was identified, and one of the records was dropped. In the rare case of triplets, three matching records were found, and two were dropped.

To account for pregnancies that did not end in a birth, we used NVSS fetal death records. In the United States, state laws require the reporting of fetal deaths, and federal law mandates national collection and publication of fetal death data.^17^ For each fetal death record, we recorded pregnancy and postpartum time in a manner identical to that produced from the birth data and recorded all pregnancy and postpartum time associated with each record that occurred in 2020.

Finally, to adjust for pregnancy or postpartum time that is cut short due to an incident of mortality, we examined all pregnancy-associated mortality events that occurred in 2019 through 2020 and adjusted our records of pregnancy and postpartum time. For each record of a pregnancy-associated mortality, we subtracted a number of months from the previously accumulated totals. The number of months subtracted depends on the time of death and the pregnancy status. For deaths occurring during pregnancy, all subsequent 12 months from the time of death–postpartum time–were removed from the exposure time. For these deaths, a matching record was assumed to occur in either the fetal death or birth records, which capture the exposure time during pregnancy. Because the death records do not include specific information about the precise timing of a given death associated with the postpartum period, assumptions about the time of death were necessary. For deaths occurring with the indicator “pregnant within 42 days,” the death was considered to have occurred at exactly 1 month postpartum, and the 11 subsequent months of postpartum time were subtracted from the analysis. For deaths occurring with the indicator “within 1 year of pregnancy,” the death was considered to have occurred at exactly 7 months postpartum, and the 5 subsequent months of postpartum time were subtracted from the analysis. Sensitivity analyses were conducted regarding this choice, and we found that the results were very insensitive to all choices ranging from 5 to 11 months. Given that results were not meaningfully different, we present 7 months here.

The tabulation of these records provided an estimate of the number of person-months spent in either the pregnancy or postpartum period during 2020. To obtain an estimate of the nonpregnant, nonpostpartum person-months, the pregnant and postpartum person-months were subtracted from the total person-months. The difference was treated as the estimate for nonpregnant, nonpostpartum person-months, given that all women would fall into one of the two mutually exclusive categories at any given point in time.

We calculated age-specific and race-specific mortality rates for pregnant/postpartum women and the nonpregnant/nonpostpartum general population. Rate ratios comparing rates of mortality among the pregnant/postpartum population and the nonpregnant/nonpostpartum population were calculated using a log Poisson regression model. 95% confidence intervals were also generated from these models. All comparisons were made for the total population in addition to subsets of the population by the age, race, and ethnicity groups previously mentioned.

All analyses were conducted in R version 4.5.0. This analysis was deemed exempt by the [BLINDED] Institutional Review Board.

## Results

A total of 11,177,577 pregnancies, captured through birth and fetal death records, were observed in the years 2019 through 2021. Of these, 9,905,908 pregnancies contributed person-time in 2020, summing to 6,431,384 person-years. Nonpregnant, nonpostpartum person-time totaled 69,046,070 person-years. Person-time was disproportionately high for the pregnant, postpartum population within the age range of 25 to 34. Across the study population, 2,856 deaths were observed, with 185 and 2,671 occurring among the pregnant/postpartum and nonpregnant/nonpostpartum population, respectively. A full breakdown of person-years and deaths observed in this analysis by age and race/ethnicity is shown in Table 1.

**Table 1.** Person-Years, Homicide Death Counts, and Crude Homicide Rates by Age and Race/Ethnicity for Pregnant/Postpartum and Nonpregnant/Nonpostpartum Individuals.

|  | Pregnant/Postpartum |  |  | Nonpregnant/Nonpostpartum |  |  |
| --- | --- | --- | --- | --- | --- | --- |
|  | Person-Years | Homicides | Crude Rate | Person-Years | Homicides | Crude Rate |
| Total | 6,431,384 | 185 | 2.88<br>(2.49-3.32) | 69,046,070 | 2,671 | 3.87<br>(3.72-4.02) |
| Age: |  |  |  |  |  |  |
| 10-19 | 288,256 | 21 | 7.29<br>(4.75-11.17) | 20,830,306 | 397 | 1.91<br>(1.73-2.10) |
| 20-24 | 1,176,197 | 64 | 5.44<br>(4.26-6.95) | 9,348,761 | 482 | 5.16<br>(4.72-5.64) |
| 25-29 | 1,827,876 | 41 | 2.24<br>(1.65-3.05) | 9,394,954 | 515 | 5.48<br>(5.03-5.98) |
| 30-34 | 1,914,231 | 35 | 1.83<br>(1.31-2.55) | 9,388,936 | 461 | 4.91<br>(4.48-5.38) |
| 35-44 | 1,224,824 | 24 | 1.96<br>(1.31-2.92) | 20,083,113 | 816 | 4.06<br>(3.79-4.35) |
| Race/<br>Ethnicity: |  |  |  |  |  |  |
| NH White | 2,801,889 | 58 | 2.07<br>(1.60-2.68) | 39,464,136 | 935 | 2.37<br>(2.22-2.53) |
| NH Black | 845,827 | 103 | 12.18<br>(10.04-14.77) | 10,733,668 | 1,189 | 11.08<br>(10.47-11.73) |
| Hispanic | 2,448,828 | 21 | 0.86<br>(0.56-1.32) | 14,201,019 | 472 | 3.32<br>(3.04-3.64) |

The crude homicide rate for nonpregnant/nonpostpartum women was 3.87 deaths per 100,000 (95% CI: 3.72-4.02). The crude pregnancy-associated homicide rate was 2.88 deaths per 100,000 (95% CI: 2.49-3.32). Crude homicide rates for pregnant/postpartum women exceeded the rates of nonpregnant/nonpostpartum women for ages 10-24, while crude rates were higher for nonpregnant/nonpostpartum women aged 25-44. Amongst the NH White and Hispanic populations, crude homicide rates were lower for the pregnant/postpartum women, 2.07 (1.60-2.68) and 0.86 (0.56-1.32) respectively, compared to nonpregnant/nonpostpartum, 2.37 (2.22-2.53) and 3.32 (3.04-3.64) respectively.

Across the whole study population, the risk of mortality was 35% higher among nonpregnant/nonpostpartum women compared with pregnant/postpartum women (homicide rate ratio 0.74; 95% CI 0.64-0.86). Risk of mortality was not significantly different among pregnant/postpartum women compared with nonpregnant/nonpostpartum women for either the NH White population (homicide rate ratio 0.87; 95% CI 0.66-1.13) or the NH Black population (homicide rate ratio 1.10; 95% CI 0.89-1.34). For the Hispanic population, the risk of mortality was 388% higher among nonpregnant/nonpostpartum women compared with pregnant/postpartum women (homicide rate ratio 0.26; 95% CI 0.16-0.39).

For those aged 10-19, risk of mortality was 282% higher among pregnant/postpartum women compared with nonpregnant/nonpostpartum women (homicide rate ratio 3.82; 95% CI 2.39-5.77). No significant difference was found for mortality rates between pregnant/postpartum women compared with nonpregnant/nonpostpartum for those aged 20-24. For age groups 25-44, the risk of mortality was higher among nonpregnant/nonpostpartum women compared with pregnant/postpartum women (Figure 2).

**Figure 2.**
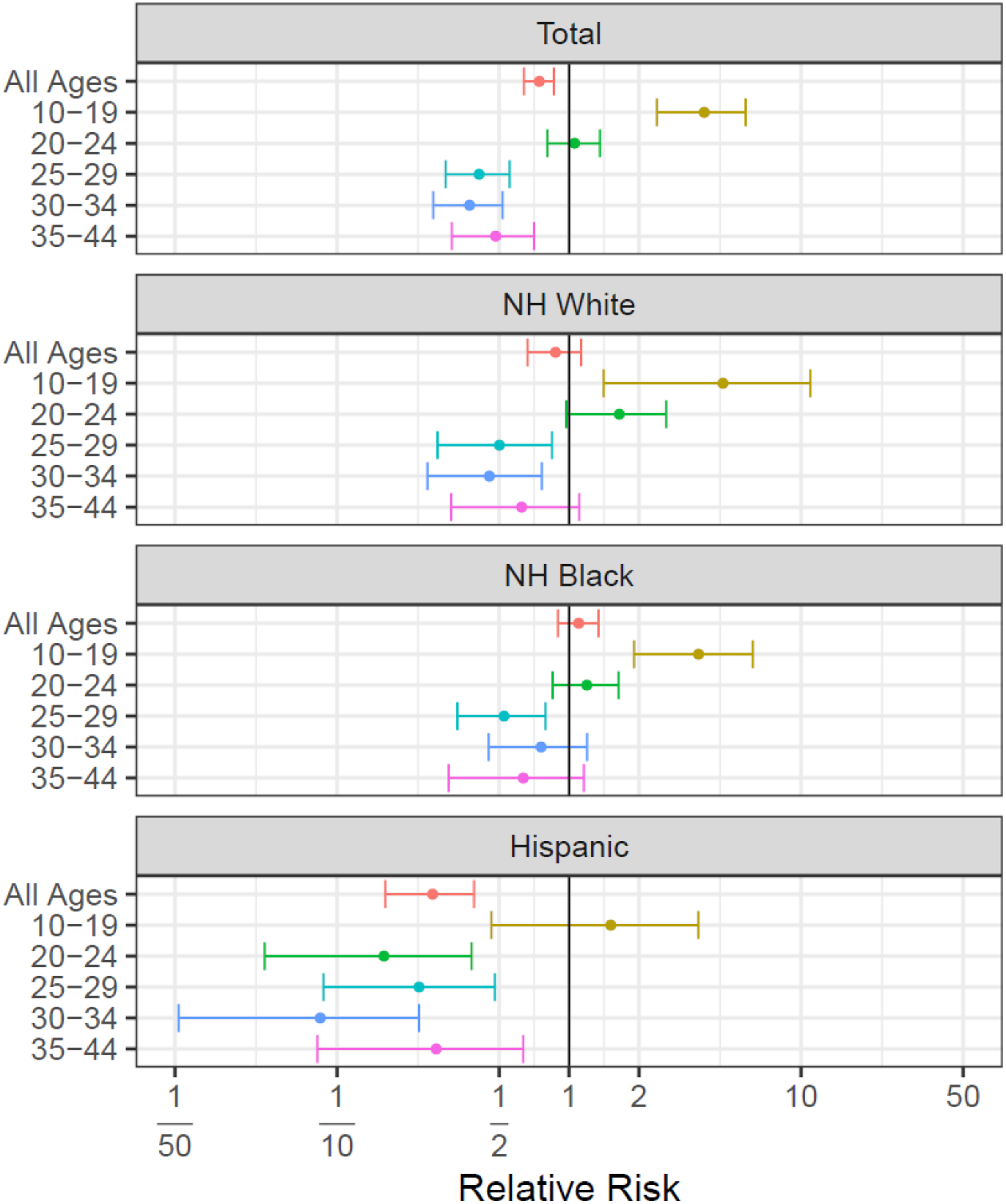
Rate ratios of homicide mortality by age and race/ethnicity. Note: Values greater than 1 indicate that a greater homicide mortality risk is present for pregnant/postpartum women compared to nonpregnant/nonpostpartum women.

## Discussion

The national expansion of a pregnancy-associated checkbox on death certificates has enabled researchers to examine pregnancy-associated mortality rates^.18^ Recent, widely circulated literature has subsequently documented a concerning and growing risk of pregnancy-associated homicide among pregnant/postpartum women compared to their nonpregnant/nonpostpartum counterparts^.9,15,19^ Existing literature on this topic, however, utilized methodologies that mischaracterized the population at risk of pregnancy-associated mortality. While previously published estimates suggest that pregnancy increases risk of homicide for all populations,^9^ our estimation process highlights that the elevated risk is only present for young populations, individuals aged 10-19. Our estimates differ because they reflect the exposure time associated with pregnancy-associated mortality. Each pregnancy event has a variable duration of exposure time associated with it, with pregnancy events on average contributing 22 months of exposure during the combined pregnancy and postpartum period. By accounting for exposure time, a more accurate comparison can be made between the nonpregnant/nonpostpartum and the pregnant/postpartum populations.

The findings of our study largely align with a paper by Dietz et al., which, back in 1998, used a similar methodology to assess injury and homicide deaths among postpartum women and their nonpostpartum peers in Georgia^.20^ In their study, only the 12-month postpartum period was compared against the nonpregnant/nonpostpartum population. By solely focusing on this 12-month postpartum period, a better comparison can be made between populations because a single birth is roughly equivalent to 12 person-months or 1 person-year of exposure time. When including the pregnancy period in addition to the 12-month postpartum period in an analysis, the denominator requires adjustments to take into account the additional exposure time of the pregnancy period.

Although we find that nonpregnant/nonpostpartum individuals are generally at higher risk of homicide compared to pregnant/postpartum individuals, race- and age-specific estimates demonstrate that the exception to this finding is among 10-19 year old pregnant/postpartum individuals, who are at heightened risk of homicide compared to their nonpregnant/nonpostpartum peers. The possible reasons for this increased risk among youth are numerous. As stated by Wallace (2022), the ability to control pregnancy status may, to some degree, have implications for the risk of homicide^.12,21^ Because youth and young adults may not have full independence and autonomy when it comes to healthcare access, restrictions to contraceptives and abortion (both related to and unrelated to the Covid-19 pandemic) may contribute to their heightened risk of pregnancy-associated homicide^.22^ Systemic approaches to improving the health and safety of pregnant and postpartum women should be tailored to particularly vulnerable populations such as pregnant and postpartum youth.^23^

In spite of our efforts to improve the standard methodology used to estimate pregnancy-associated mortality risk, our estimates reflect a few data limitations. First, while the population-based data used here are a strength of our analysis, our estimates are limited by small cell sizes, given the relative rarity of the outcomes (i.e., pregnancy-associated mortality). Second, the pregnancy checkbox is known to include false-positives and false-negatives^.24^ For example, pregnancy status may not be known for individuals who are early in the first trimester. For pregnancies that do not end in a birth, the date of the termination may be unknown or uncertain. Because pregnancy-associated deaths are relatively rare, it has not been well established how misclassification might differ across subgroups; future research should continue to examine the impact of the pregnancy checkbox and its potential flaws.^7^ Finally, in this paper we focus on homicide risk; however, future work should apply our methodology to other causes of death.

## Public Health Implications

Although our findings differ from previously published estimates, the implications remain clear: policies and strategies that address social and environmental risk to mitigate homicide are urgently needed. However, the development of effective policies and strategies relies on the accurate assessment of pregnancy-associated mortality risk. Our findings suggest that policies addressing homicide risk among pregnant and postpartum individuals might be particularly effective if they are focused on the protection of young people. Future work on this topic should use a more accurate denominator to ensure estimates reflect the true burden of mortality among pregnant and postpartum women overall and among specific vulnerable subpopulations.

## Data Availability

All individual, restricted-use data must be obtained from the National Vital Statistics System directly. An aggregated research dataset can be made available upon reasonable request to the authors.

https://www.cdc.gov/nchs/nvss/nvss-restricted-data.htm

## References

1. Creanga AA, Berg CJ, Syverson C, Seed K, Bruce FC, Callaghan WM. Pregnancy-Related Mortality in the United States, 2006–2010. Obstetrics & Gynecology. 2015;125(1):5. doi:10.1097/AOG.0000000000000564

2. Collier AY, Molina RL. Maternal Mortality in the United States: Updates on Trends, Causes, and Solutions. NeoReviews. 2019;20(10):e561–e574. doi:10.1542/neo.20-10-e561

3. MacDorman MF, Declercq E, Thoma ME. Trends in Maternal Mortality by Sociodemographic Characteristics and Cause of Death in 27 States and the District of Columbia. Obstetrics & Gynecology. 2017;129(5):811–818. doi:10.1097/AOG.0000000000001968

4. Creanga AA, Berg CJ, Ko JY, et al. Maternal Mortality and Morbidity in the United States: Where Are We Now? J Womens Health (Larchmt). 2014;23(1):3–9. doi:10.1089/jwh.2013.4617

5. National Center for Health Statistics. Maternal Mortality FAQ. cdc.gov. April 29, 2024. Accessed October 5, 2025. https://www.cdc.gov/nchs/maternal-mortality/faq.htm

6. CDC. Reference Guide for Pregnancy-Associated Death Identification. Maternal Mortality Prevention. May 20, 2024. Accessed August 28, 2025. https://www.cdc.gov/maternalmortality/php/mmrc/reference-guide.html

7. Rossen LM, Womack LS, Hoyert DL, Anderson RN, Uddin SFG. The Impact of the Pregnancy Checkbox and Misclassification on Maternal Mortality Trends in the United States, 1999-2017. Vital Health Stat 3. 2020;(44):1–61.

8. Cliffe C, Miele M, Reid S. Homicide in pregnant and postpartum women worldwide: a review of the literature. J Public Health Pol. 2019;40(2):180–216. doi:10.1057/s41271-018-0150-z

9. Wallace M, Gillispie-Bell V, Cruz K, Davis K, Vilda D. Homicide During Pregnancy and the Postpartum Period in the United States, 2018–2019. Obstetrics & Gynecology. 2021;138(5):762. doi:10.1097/AOG.0000000000004567

10. Wallace ME, Jahn JL. Pregnancy-Associated Mortality Due to Homicide, Suicide, and Drug Overdose. JAMA Netw Open. 2025;8(2):e2459342. doi:10.1001/jamanetworkopen.2024.59342

11. Dholakia A, Monuteaux MC, D’Ambrosi G, McLone SG, Fleegler E, Lee LK. Firearm Homicide in Pregnant Women and State-Level Firearm Ownership. JAMA Netw Open. 2025;8(11):e2542447. doi:10.1001/jamanetworkopen.2025.42447

12. Wallace ME. Trends in Pregnancy-Associated Homicide, United States, 2020. Am J Public Health. 2022;112(9):1333–1336. doi:10.2105/AJPH.2022.306937

13. Wallace ME, Hoyert D, Williams C, Mendola P. Pregnancy-associated homicide and suicide in 37 US states with enhanced pregnancy surveillance. American Journal of Obstetrics and Gynecology. 2016;215(3):364.e1-364.e10. doi:10.1016/j.ajog.2016.03.040

14. Lesko CR, Fox MP, Edwards JK. A Framework for Descriptive Epidemiology. American Journal of Epidemiology. 2022;191(12):2063–2070. doi:10.1093/aje/kwac115

15. Wallace M. Trends in Pregnancy-Associated Homicide, United States, 2020. Am J Public Health. 2022;112(9):1333–1336. doi:10.2105/AJPH.2022.306937

16. ICD-10 Version:2019. Accessed February 4, 2024. https://icd.who.int/browse10/2019/en

17. National Vital Statistics System. Fetal Deaths. CDC National Center for Health Statistics. March 18, 2025. Accessed September 29, 2025. https://www.cdc.gov/nchs/nvss/fetal_death.htm

18. Rossen LM, Womack LS, Hoyert DL, Anderson RN, Uddin SFG. The Impact of the Pregnancy Checkbox and Misclassification on Maternal Mortality Trends in the United States, 1999-2017. Vital Health Stat 3. 2020;(44):1–61.

19. Wallace ME, Jahn JL. Pregnancy-Associated Mortality Due to Homicide, Suicide, and Drug Overdose. JAMA Network Open. 2025;8(2):e2459342. doi:10.1001/jamanetworkopen.2024.59342

20. Dietz PM, Rochat RW, Thompson BL, Berg CJ, Griffin GW. Differences in the risk of homicide and other fatal injuries between postpartum women and other women of childbearing age: implications for prevention. Am J Public Health. 1998;88(4):641–643.

21. Vilda D, Wallace ME, Daniel C, Evans MG, Stoecker C, Theall KP. State Abortion Policies and Maternal Death in the United States, 2015-2018. Am J Public Health. 2021;111(9):1696–1704. doi:10.2105/AJPH.2021.306396

22. Bryson AE, Hassan A, Goldberg J, Moayedi G, Koyama A. Call to Action: Healthcare Providers Must Speak Up for Adolescent Abortion Access. Journal of Adolescent Health. 2022;70(2):189–191. doi:10.1016/j.jadohealth.2021.11.010

23. Wollum A, Harmon S, Thompson TA. Do remote judicial bypass hearings increase access for minors seeking abortion care?: A quasi-experimental study, 2018–2023. Contraception. 2026;153:111245. doi:10.1016/j.contraception.2025.111245

24. Catalano A, Davis NL, Petersen EE, et al. Pregnant? Validity of the pregnancy checkbox on death certificates in four states, and characteristics associated with pregnancy checkbox errors. Am J Obstet Gynecol. 2020;222(3):269.e1-269.e8. doi:10.1016/j.ajog.2019.10.005

